# Operational Enablers and Barriers in Hospital Incident Command: Insights from a Single-Center Table-Top Exercise at a Tertiary Care University Hospital—A Qualitative Phenomenological Study

**DOI:** 10.64898/2026.05.13.26353139

**Authors:** Markus Ries, Maik von der Forst, Hanne Schäfer, Kirsten Bikowski, Klaas Franzen, Paul Geoerg, Fabian Weykamp, Erik Popp, Janna Küllenberg

## Abstract

**Background:** In crises, hospitals must rapidly shift from routine operations to structured crisis management, requiring the activation of an incident command system. However, empirical insight into their operational functioning during activation remains limited. Goal: to identify operational enablers and barriers influencing effective crisis response.

**Methods:** Prospective cross-sectional, qualitative, single-center study conducted after a table-top exercise within a hospital incident command system at a tertiary care university hospital (NCT06913010). Data was collected through semi-structured interviews, participant observation, and document analysis, and analyzed using a narrative-phenomenological approach.

**Results:** Nineteen participants were included. Analysis identified nine thematic clusters shaping operational performance: (1) structure and roles; (2) communication; (3) decision-making and prioritization; (4) information management; (5) infrastructure and technology; (6) personnel and organization; (7) training, exercises, and team dynamics; (8) documentation; and (9) external communication and media. Enablers included clear role definition, structured communication, phased decision-making, and regular training. Barriers included role ambiguity, fragmented communication, insufficient prioritization, infrastructure limitations, and staffing constraints.

**Conclusion:** Preparedness frameworks are necessary but insufficient as stand-alone approaches, as operational execution determines real-world performance. Recurring deficits included unclear roles, inconsistent communication, weak prioritization, and gaps in infrastructure and personnel. A limited set of standardized practices - including a clear separation od roles, leadership intent, closed-loop communication, explicit decision cycles from information gathering to structuring to decision-making, checklists, visualization, central information management, and rapid “80% decisions”-substantially enhanced performance. Mission command (*Auftragstaktik)* further enabled adaptive, coordinated action. Strengthening hospital incident command is a key lever for achieving system-level resilience in crises.

## Introduction

Health security encompasses the capacity of health systems to prevent, detect, and respond to public health threats and complex emergencies (1,2). In this context, hospitals play a central role as providers of acute care and as key actors in maintaining healthcare system functionality during crises. Large university hospitals represent critical infrastructure, as their continuous operation is essential for population health and system stability (3). Disruptions - whether due to internal failures or external hazards - can have cascading effects on regional healthcare delivery and emergency response systems (4,5). Hospitals are exposed to a wide range of potential threat scenarios, including cyberattacks, infectious disease outbreaks, mass casualty incidents, infrastructure failures, and other complex multi-hazard events (3,4,6,7). Such VUCA (Volatility, Uncertainty, Complexity, Ambiguity) situations are typically characterized by uncertainty, time pressure, incomplete information, and the need for rapid coordination across multiple actors (8). To manage these challenges, hospitals are required to transition from routine clinical operations to a structured crisis management mode. This transition is commonly operationalized through the activation of a hospital incident command system (HICS), which establishes a dedicated leadership and coordination structure (9,10). In Germany, this structure is frequently aligned with the continental staff system as described in operational guidelines such as *DV 100* (German Fire Service regulation “*Dienstvorschrift 100”*, outlining command and control structures in emergency response), providing a framework for coordinated decision-making and information processing in complex situations (11–14).

The implementation and training of hospital emergency management structures are embedded within a multi-level regulatory architecture. The World Health Organization (WHO) has developed instruments such as the International Health Regulations (IHR), the IHR Monitoring and Evaluation Framework, and operational tools including the Hospital Readiness Checklist, and the Hospital emergency response checklist which are used globally to guide hospitals and health systems in preparing for and responding to health emergencies (15–18). At the national level in Germany, the Federal Office of Civil Protection and Disaster Assistance (in German: *BBK)* provides structured guidance for the development of hospital emergency and response plans (in German: *KAEP*). To support consistent capability development across institutions, hospitals have the option to obtain certification through the German Association for Hospital Emergency Planning (in German: DAKEP) as the relevant professional society. (19–21). These requirements are further specified regionally within the German healthcare system, for example through the framework for hospital emergency and response management in the federal state of Baden-Württemberg (22). Across these levels, regular training and exercises including, but not limited to the incident command system, are recommended to ensure operational readiness and to develop the necessary competencies for crisis response (23).

Despite these established frameworks, the scientific evidence on the effectiveness of hospital emergency preparedness training remains limited, and there is a particular lack of empirical insight into how HICS function under operational conditions (24–28). Previous research has demonstrated that structured training and large-scale simulation exercises can improve staff knowledge, role clarity, and familiarity with emergency procedures, and may enhance preparedness at the organizational level (29,30). Complementing this line of research, the present study focuses on a table-top exercise, representing the smaller-scale end of the exercise spectrum, and examines cognitive, communicative, and decision-making processes within the incident command structure rather than full operational execution. Against this background, a structured training and exercise program for a staff-based HICS was conducted at a large university hospital. The aim of this exercise was to prepare key personnel for crisis situations and to enable systematic evaluation of operational processes, communication, and leadership dynamics during the transition from routine operations to emergency mode (13).

This study is part of a prospectively registered interdisciplinary qualitative research project, applying a constructivist Grounded Theory approach to triangulated data (ClinicalTrials.gov NCT06913010) (13,31). While the overarching project aims to develop a theoretical model of the *leadership orchestration* process (manuscript in preparation), the present report presents an additional secondary, narrative-phenomenological analysis focusing on *operational enablers and barriers*. The separation into two reports was undertaken to ensure analytical depth and to provide distinct contributions - one conceptual and one applied - while maintaining transparency through cross-referencing.

We therefore directed our efforts toward identifying operational factors that influence the effectiveness of crisis management within a HICS, using a table-top exercise as a model use case. To guide this analysis, the following research questions were formulated:

Research question 1: What operational factors *facilitate* effective crisis management within a hospital incident command system, as demonstrated in a table-top exercise?

Research question 2: What operational factors *hinder* effective crisis management within a hospital incident command system, as demonstrated in a table-top exercise?

This was in alignment with the predefined primary objective of the overall research project to ‘assess training and exercise effects within a structured HICS including challenges, resistance, benefits, subjective participant experiences, potential improvements, and alternatives,’ as well as the secondary objectives to evaluate ‘participants’ operational confidence,’ ‘communication behaviors within the command functions,’ and the ‘advantages and disadvantages of a structured hospital incident command system’(31).

## Methods

### Study type and design

This study forms part of a prospectively registered interdisciplinary research program examining crisis management processes in hospital incident command structures, centered on a structured table-top exercise of a staff-based HICS. This additional, secondary analysis focuses on operational factors and was designed as a qualitative, single-site investigation using a narrative-phenomenological approach as described by Creswell (32,33). The primary analysis of this dataset applied a constructivist grounded theory framework, (13,34,35) and is reported in a separate manuscript (in preparation)

### Research checklists

To ensure scientific rigor, design, conduct, analysis, and reporting of this study followed established methodological standards, including the Standards for Reporting Qualitative Research (SRQR) and the STrengthening the Reporting of OBservational Studies in Epidemiology (STROBE) guidelines (36,37).

### Setting, training, exercise design and allocation of participants

The study was conducted at a single-center tertiary care university hospital and was embedded in a planned table-top exercise, focusing on the activation and performance of the hospital incident command structure. Prior to the table-top exercise, a standardized training session was conducted, introducing key roles, processes, and principles of hospital incident command. Prior to the table-top exercise, a standardized training session was conducted, introducing key roles, processes, and principles of hospital incident command. The exercise was carried out on two separate training days using the same scenario and within the same institutional context, thereby addressing comparable operational challenges. Allocation of participants to the respective exercise days was determined by scheduling and individual availability. Participants were not randomized. To minimize potential information bias between the exercise days, participants in the first session were instructed to maintain strict confidentiality regarding the exercise content and procedures until completion of the second session. After completing the exercise, participants were offered enrollment in this study.

### Data sources

Data were derived in a triangulated approach analyzing semi-structured interviews, participant observation, and exercise-related documents. The main data source for this analysis was semi-structured interviews with participants who were involved in the hospital incident command training and exercise. The interviews focused on participants’ experiences, perceptions, and interpretations of operational processes, communication, and leadership dynamics during the transition from routine operations to crisis mode, with particular emphasis on operational phenomena that facilitated or hindered effective crisis management as experienced during the table-top exercise. In addition, observational data were collected during the exercise as open, non-structured participant observation, including field notes and observation sheets. Furthermore, exercise-related documents - such as protocols, photographic documentation of flipcharts, and evaluation materials generated during pre-briefing, debriefing, and feedback processes - were incorporated as supplementary data sources. Consistent with the principles of constructivist Grounded Theory, all data sources were considered complementary and contributed to a comprehensive understanding of the studied phenomena.

### Ethics and study registration

All study procedures were conducted in accordance with the Declaration of Helsinki and applicable data protection regulations, including the European Union General Data Protection Regulation, the German Federal Data Protection Act, and the State Data Protection Act of Baden-Württemberg. Germany. The study underlying the present analysis was approved by the Ethics Committee of the Medical Faculty of Heidelberg University (reference S-065/2025). Written informed consent was obtained from all participants prior to inclusion. The study was prospectively registered on ClinicalTrials.gov (NCT06913010) prior to enrollment and execution.

### Participants, inclusion and exclusion criteria

Participants were recruited from employees of a single-site, large university hospital who were eligible for deployment within the HICS. Inclusion criteria comprised adult staff members with the ability to provide informed consent who had participated in the HICS exercise. Written informed consent was required for participation. The sole exclusion criterion was refusal to provide written informed consent. Sampling followed the procedures of the overarching study, participants were interviewed, with each individual offering potential conceptual variation. This comprehensive sampling facilitated analysis and provided phenomenological sufficiency.

### Semi-structured interview guide and Interviews

The semi-structured interview guide was developed by the study team in accordance with the collect-sort-check-delete-subsume procedure (in German: S^2^PS^2^) procedure (13,38). The guide was sequentially structured along the temporal progression of the exercise to capture participants’ experiences along the temporal and operational progression of the exercise, consisting of an open narrative introduction followed by probe questions to ensure thematic coverage (13).

Semi-structured interviews were conducted with eligible participants after completion of the table-top disaster management exercise. Interviews were scheduled individually and conducted using web-based video conferencing software. All interviews were audio-recorded and transcribed in pseudonymized form using MAXQDA (VERBI Software, Berlin, Germany)

### Training and HICS table-top exercise timeline

The study followed a sequential process, with training sessions and the table-top exercise conducted as preparatory activities preceding the study, followed by participant recruitment and inclusion, and concluding with the conduct of semi-structured interviews. The training sessions were conducted on April 29, 2025, and May 20, 2025. The table-top incident command exercise took place on May 26 and 27, 2025, with a duration of 4.5 hours per session. Semi-structured interviews were conducted between May 30, 2025, and August 16, 2025. All activities were carried out at the University Hospital Heidelberg, Germany.

### Qualitative data analysis: Coding of significant statements, horizontalization, grouping into clusters of meaning

Qualitative data analysis was conducted following a narrative-phenomenological approach as described by Creswell and as applied in previous health security and public health research (23,32,33,39–41). The phenomenological approach was chosen to capture lived operational experiences, aligning with a constructivist-interpretivist paradigm that emphasizes sensemaking in dynamic and uncertain contexts. Transcripts were examined for significant statements, which were treated as the primary units of analysis with equal weighting. These statements were subsequently horizontalized and grouped into clusters of meaning to identify recurring themes and patterns across participants’ experiences. This process enabled a structured exploration combining deductively informed data collection with inductive identification of themes and patterns of operational processes, communication dynamics, and perceived enablers and barriers within the HICS.

### Variables and codes

Variables and codes were developed iteratively during the qualitative analysis. Coding was performed using the open-source qualitative data analysis software QualCoder (version 3.3, Ubuntu), running on Linux Mint 21.3 (LTS, ‘Virginia’) (42,43).

Specifically, codes and clusters of meaning variables included: age, sex, profession, years of experience in current role, significant statements reflecting operational facilitating factors, significant statements reflecting operational hindering factors, and the following clusters of meaning: (1) structure and roles, (2) communication, (3) decision-making and prioritization, (4) information management, (5) infrastructure and technology, (6) personnel and organization, (7) training, exercises, and team dynamics, (8) documentation, and (9) external communication and media. The text was coded in the German language source document transcripts, results were subsequently translated into English. A mind map was developed to support the structured visualization and organization of clusters of meaning and to facilitate the exploration of relationships and interdependencies between thematic domains. The mind map was created using Minder version 2.0.5, an open-source mind-mapping application running on inux Mint 21.3 (LTS, ‘Virginia’) (43,44).

### Techniques to enhance trustworthiness

Methodological rigor was ensured through detailed and transparent reporting of the research process, to allow transferability to comparable settings. Credibility was strengthened by the interdisciplinary composition of the research team, integrating complementary perspectives from different professional backgrounds and enabling a comprehensive 360° understanding of the study context. All team members had extensive professional experience relevant to the study topic. All core team members were embedded in the exercise as observers, providing in-depth contextual insight. In addition, external observers were involved, including individuals from the same institution as well as from other institutions, thereby contributing independent perspectives, supporting reflexivity, and enhancing the credibility and robustness of the analysis. Formal communicative validation (member checking) was not conducted, as the study was based on a time-limited exercise setting and post hoc qualitative analysis. To ensure analytical rigor and enhance dependability and confirmability, multiple strategies were applied, including triangulation of data sources, involvement of external observers, and structured, team-based reflexive analysis.

### Statistical analysis for qualitative and quantitative data

Statistical or frequency-based analyses of the codes and clusters of meaning were not conducted, as the phenomenological approach prioritizes the interpretation of meaning over quantification. In addition, the relatively small sample size (N = 19) and the broad spectrum of response themes limited the interpretability of frequency distributions. The analysis therefore focused on depth, context, and thematic relationships.

Quantitative variables were analyzed using descriptive statistical methods, including calculation of sample size (N), means and standard deviations for parametric data, medians and interquartile ranges for non-parametric data, as well as minimum and maximum values. Categorical variables were summarized using frequencies and percentages. Missing values were not imputed. Subgroup analyses were not performed. All analyses were performed using R version 4.5.3 and RStudio version 2026.01.2+418 running on Linux Mint 21.3 (LTS, ‘Virginia’) (43,45,46).

### Inductive framework development and deductive juxtaposition

An inductive analytical approach was used to allow operational patterns, enablers, and barriers to emerge directly from the data without imposing predefined categories. This was considered more open and appropriate given the complex and partially under-theorized nature of hospital incident command processes in simulated crisis settings. In the discussion, the inductively derived framework was juxtaposed with established resilience and emergency management frameworks to enable a theory-informed, deductive comparison of areas of convergence and divergence.

### Researcher characteristics and reflexivity

Qualitative data analysis and translation processes may be influenced by the characteristics, professional backgrounds, and reflexivity of the researchers involved. To address this, the study team was deliberately composed to ensure methodological rigor while incorporating diverse perspectives relevant to disaster medicine and hospital operations. Translations were performed by a member of the research team and were carefully reviewed to ensure semantic and contextual equivalence. Selected excerpts were back-translated as an internal validation step. The analysis group consisted of five embedded and three non-embedded researchers with complementary expertise: (1) a social education professional with expertise in communication and employee guidance and no prior disaster exercise experience; (2) a psychologist with strong qualitative methodological expertise and limited disaster exercise experience; (3) a physician-scientist with international disaster medicine certification and a civil–military background; (4) the head of operative hospital emergency and response planning with extensive experience in disaster exercises and voluntary disaster management; and (5) a physician with extensive hospital emergency management and clinical operations experience. This composition enabled both insider and outsider perspectives on the data. To enhance analytical rigor and credibility, three external validators independent of the table-top exercise contributed to the interpretation process: (6) a radiation oncologist with clinical and research expertise in interdisciplinary care and radiation emergency preparedness and response (local), (7) a physician with extensive experience in hospital emergency management and disaster medicine (external to the study site), and (8) a researcher specializing in complex system dynamics, including civic safety, pedestrian and crowd dynamics and data engineering in civil safety contexts (external to the study site), thereby supporting reflexive interpretation and mitigating potential bias arising from the embedded research context.

### Positionality statement

The research team occupies a position of structural proximity to the phenomenon under study: several embedded researchers hold or have held operational roles within the HICS at the study site, and the exercise itself was designed and conducted by members of the same institutional unit. This insider positionality carries the epistemological risk that operational assumptions, institutional preferences, or professional blind spots may have shaped the coding of significant statements and the formation of thematic clusters in ways that are difficult to fully make transparent.

To address this, the analytical process was deliberately structured to foreground disconfirming evidence. Non-embedded researchers and external validators were specifically asked to challenge emerging interpretations and to identify phenomena that might have been underweighted by the embedded team. Analytical memos documenting points of disagreement and resolution were maintained throughout the process. Additionally, the use of verbatim participant quotes as the primary unit of analysis - rather than researcher-constructed summaries - provided an anchoring mechanism that reduced the scope for post-hoc interpretive drift.

Readers should nonetheless interpret the findings with awareness of this structural positionality and consider the degree to which the identified enablers and barriers may reflect not only participants’ lived experiences but also the conceptual frameworks that are institutionally dominant at the study site.

### AI use statement

The authors used artificial intelligence (AI) tools (Chat GPT, OpenAI) to assist with language editing, as all authors are non-native English speakers. AI use was rigorously human-centered, and no AI tools were used for data analysis or interpretation. All outputs were carefully reviewed, validated, and revised by the authors, who take full responsibility for the accuracy, integrity, and originality of the manuscript.

## Results

### Population sample

Twenty-three individuals participated in the HICS exercise and were assessed for eligibility. Of these, 20 consented to participate; one participant dropped out due to scheduling conflicts, and the remaining 19 completed the interviews.

Participants represented a highly interdisciplinary cohort spanning senior clinical, technical, administrative, and executive leadership roles, and only one person without contact to HICS before conducting the actual exercise (**Table 1**).

**Table 1.** Demographic and interdisciplinary professional characteristics of study participants (N=19) in the single-center hospital incident command table-top exercise

| Characteristic | Data |
| --- | --- |
| Sex | Female: 7 (37%); Male: 12 (63%) |
| Participation in the table-top HICS exercise | Yes: 19 (100%) |
| Age (years) | Mean 50.4, SD 9.37, Median 55, IQR [43...58], Range 33–61, N = 19 |
| Experience in current role (years) | Mean 5.5, SD 6.90, Median 3, IQR [2...6], Range 0.67–29, N = 19 |
| **Professional background** | **Clinical/Healthcare (N=6):** Nursing management; Physician (N=4); Dentist.<br>**Technical/Engineering/IT (N=7):** IT specialist; Graduate engineer; Mechanical engineer (Dipl.-Ing.); HVAC master craftsman/Business administrator; Automotive mechatronics technician; Computer scientist; Industrial engineer. **Administration/Management/Strategy (N=3):** Lawyer; Diploma in Nursing Management; PR Consultant (B.Sc. Chemistry).<br>**Communication/Public Interface (N=1):** Press spokesperson.<br>**Safety/Operations/Support (N=2):** Specialist for protection and security; Cook. |

HICS = hospital incident comment system

The analysis identified a set of operational hindering and facilitating factors across nine thematic clusters that collectively shaped the effectiveness of crisis response. These clusters are illustrated in **Figure 1** and are then described in detail in the following sections. Each cluster is structured into (1) facilitating factors, (2) an illustrative “power quote,” (3) hindering factors, and (4) a concise summary of the key insights. Interview excerpts are presented using anonymized interview identifiers (e.g., I1–I12) to ensure participant confidentiality.

**Figure 1.**
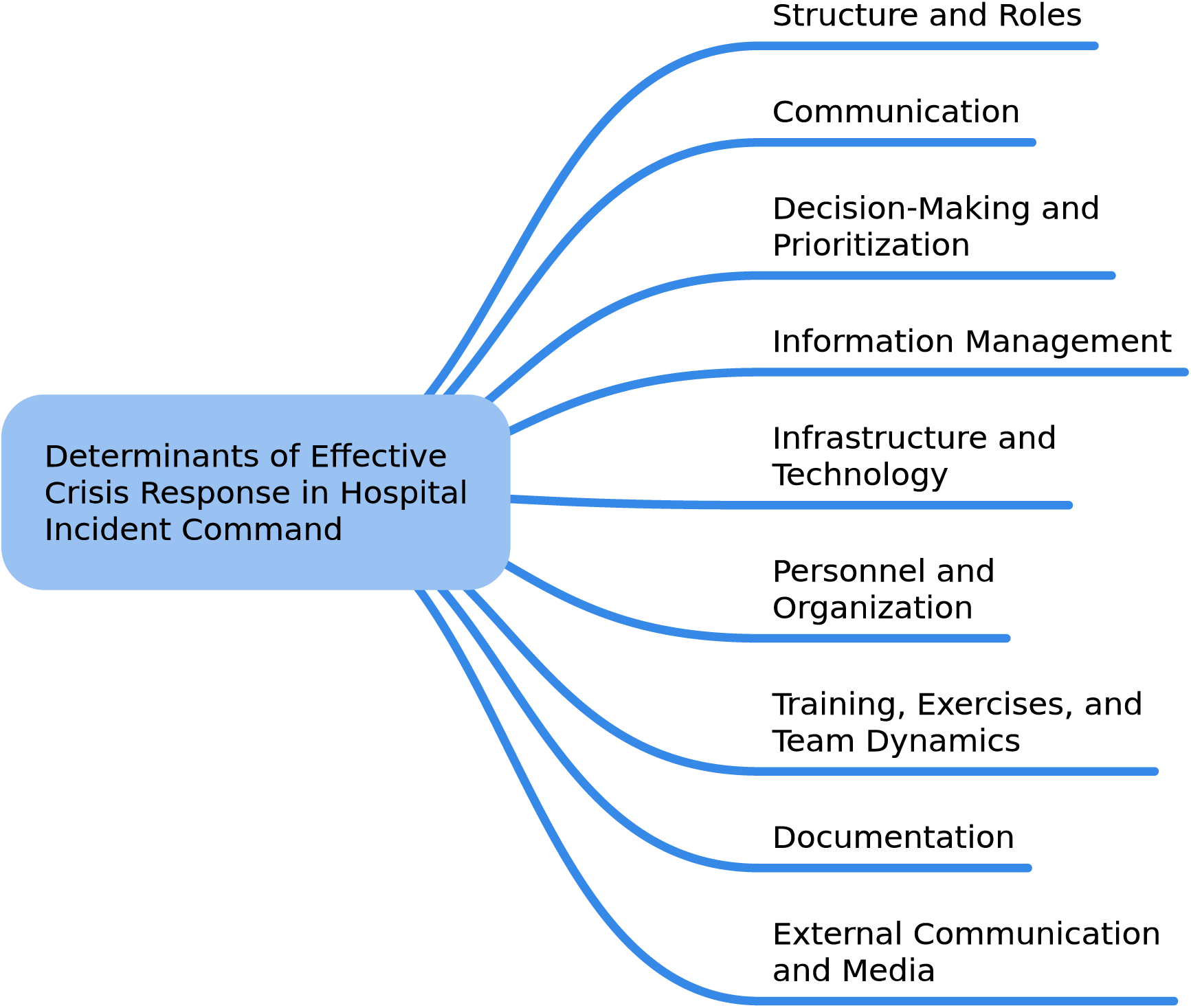
Mind map illustrating nine thematic clusters derived from significant statements of the study participants (N=19) on enabling and hindering factors shaping determinants of effective crisis response within a hospital incident command system. The clusters do not imply hierarchy or sequencing.

#### 1. Structure and Roles

Structure and roles emerged as a central domain shaping coordination and decision clarity. Enabling factors included a clear definition of roles within and across leadership, facilitation, and documentation, predefined substitution arrangements, standardized role profiles and standard operating procedures, and regular briefings on roles and authorities. It was also considered important that roles and individuals could be clearly and visually identified within the team. One participant noted, “I think that what was important was that you knew that everyone had their name on their jacket. I think it is extremely important that you can identify people, if you know what function they have, even without knowing their name” (I5). Hindering factors included missing or unclear role allocation, dual roles, and uncertainty regarding decision responsibilities. In addition, participants often remained in their routine roles instead of transitioning into designated crisis roles, which limited effective coordination. Overall, clear role definition and separation of responsibilities enhanced coordination and decision-making, whereas role ambiguity, dual functions, and failure to transition into crisis roles undermined effective operational performance.

#### 2. Communication

Communication was pivotal for maintaining shared situational awareness and coordinated action. Enabling factors included short, clear, and precise communication in line with crew resource management principles, the use of predefined communication pathways without parallel side channels, and regular status updates and structured check-ins. One participant emphasized, “Clarity in communication, so that the counterpart - whoever it may be - knows what the full scope is, what the problem is, what solution pathways we have, what tasks we have, and who is doing what” (I10). In addition, the explicit labeling of sensitive information and the establishment of a crisis-resilient internal communication system with defined fallback layers (e.g., radio communication, designated runners) supported information flow and coordination. Hindering factors included chaotic communication and misunderstandings, missing or delayed feedback, the use of side channels and unfiltered external communication, unclear communication pathways between levels, and incomplete or inconsistent information within the situational picture. Taken together, structured and disciplined communication improved situational awareness and coordination, whereas fragmented communication, unclear pathways, and incomplete information compromised effective crisis management.

#### 3. Decision-Making and Prioritization

Decision-making and prioritization defined the timeliness and direction of operational responses. Enabling factors included prioritization according to the principle of “life and safety first,” as one participant pointed out, “You always first look at whether life and limb are at risk” (I13). Equally important is a clear separation of phases into (1) information gathering, (2) structuring, and (3) decision-making, as well as the use of checklists to support prioritization. In addition, the acceptance of “80% decisions” (i.e., decisions made on sufficient but incomplete information to prioritize timeliness over full optimization) to enable timely action as well as the visualization of tasks, deadlines, and responsibilities facilitated structured and efficient decision-making, as one participant reflected: “A best-possible decision can also be an […] 80% decision that is made twice as fast as making a [fully optimized (added for clarity)] decision” (I12). Hindering factors included a lack of prioritization of immediate actions, time pressure leading to the premature termination of important discussions, the conflation of information gathering and decision-making processes, and uncertainty regarding urgency. In summary, structured prioritization and clear decision-making processes improved timeliness and effectiveness, whereas unclear priorities, time pressure, and unstructured processes impaired operational performance.

#### 4. Information Management

Information management underpinned the development of a coherent and actionable situational picture. Enabling factors included the use of a structured situational overview matrix distinguishing between known information, unknown information, and required actions. One participant described: “Typically, you first document the measures as well as the known and unknown conditions.” (I16). In addition, the establishment of a central collection point for information (e.g., back office or command support staff) further supported effective information management. Likewise, the use of visualization tools such as flipcharts or software supported the organization and communication of information. The introduction of a standardized operational logbook improved documentation and traceability. Hindering factors included incomplete and partly contradictory situational awareness, unsystematic information gathering, missing feedback from operational units or departments, and additional information that was not always relevant or delayed. Altogether, structured information management and centralized processing improved situational clarity and coordination, whereas fragmented, delayed, or inconsistent information diminished effective decision-making and operational performance.

#### 5. Infrastructure and Technology

Infrastructure and technology provided the foundation for system resilience and response capability. Enabling factors included investments in redundancy and resilience (e.g., backup power supply and backup IT systems), the establishment of crisis containers or emergency carts, and the implementation of standardized emergency IT concepts and backup systems. In addition, clear coordination agreements with external partners such as the Federal Agency for Technical Relief (in German: THW), fire services, and suppliers, as well as regular technical testing and failure simulations, supported system reliability and preparedness. As one participant reflected based on prior experience, “I’m someone who strongly believes that we need to practice our emergency concepts from time to time. […] There are hospitals that deliberately shut systems down in a controlled way […] because when something like that almost never fails, I can tell you—at first, nobody knows what to do. […] And we experienced this a few years ago. It really showed us that you first must actively bring these emergency concepts into practice” (I10). Hindering factors included partly outdated infrastructure, a lack of redundancies in power supply, IT, and equipment, insufficient technical equipment (e.g., laptops, service phones, radio systems), vulnerabilities in IT systems, and unclear arrangements regarding emergency power supply and fuel provision. Overall, robust and redundant infrastructure enhanced operational resilience, whereas outdated systems, insufficient equipment, and unclear contingency arrangements limited effective crisis response.

#### 6. Personnel and Organization

Personnel and organizational factors determined system readiness and functional capacity under stress. Enabling factors included regular training and short refresher sessions, the involvement of pre-designated decentralized disaster preparedness officers, the establishment of a structured crisis readiness system, and the promotion of psychological safety and a constructive error culture. As one participant noted, “There was nothing that could not be said” (I11), while another acknowledged, “Mistakes happen, misunderstandings occur - I believe that will always be the case in some way.” (I15). Hindering factors included staff shortages, particularly in nursing and technical services, insufficient training for crisis situations, limited ad hoc availability of personnel, and knowledge gaps among newly recruited staff. In summary, well-trained and readily available personnel supported effective crisis response, whereas staffing shortages, insufficient training, and organizational gaps reduced operational capacity.

#### 7. Training, Exercises, and Team Dynamics

Training, exercises, and team dynamics shaped team performance and adaptive capacity. Enabling factors included regular exercises, including unannounced scenarios, the integration of realistic stress situations and dilemmas, and the involvement of all organizational levels (clinical departments, technical services, and administration). As one participant suggested, “In addition to the structures we are currently establishing, it is important to run through things regularly, at defined intervals. […] Not just as a pure hospital incident command exercise, but also by involving the clinical areas” (I10). In addition, improved preparation of new members and implementation of structured debriefings supported team development and learning. Hindering factors included a hesitant start of the exercise, delayed team formation, the artificial nature of the exercise setting with a lack of realistic stressors, and declining concentration during prolonged sessions. Taken together, realistic and regularly conducted training strengthened team cohesion and preparedness, whereas artificial settings, delayed team dynamics, and fatigue limited the effectiveness of exercises.

#### 8. Documentation

Transparent documentation ensured traceability of decisions and actions. Enabling factors included the assignment of a dedicated person responsible for documentation, the use of visible real-time documentation tools (e.g., software or whiteboards capturing “who does what and by when”), mandatory feedback on task completion, and structured documentation during each situation briefing. As one participant illustrated, “I would display it using a projector, if possible - put it up on the wall – bang - and then ask precisely: where do you stand? What is holding things up?” (I16). Hindering factors included unclear documentation roles, incomplete records of measures and tasks, and the absence of a consistently maintained operational logbook. Altogether, clear responsibilities and structured, real-time documentation improved transparency and coordination, whereas unclear roles leading to incomplete records eroded traceability and operational effectiveness.

#### 9. External Communication and Media

External communication and media management secured message consistency and public credibility. Enabling factors included the centralized handling of all external information through the press office, the implementation of standard operating procedures for social media and media inquiries, the establishment of crisis websites or emergency information channels, and the involvement of external expertise in crisis communication. One participant explained, “It is then entirely clear that something has to be drafted, that it goes through some form of approval process, and that it is then released” (I14). Hindering factors included the premature release of internal information to external audiences, inadequate integration of the press office, and unclear strategies for social media communication. Overall, centralized and structured external communication strengthened message control and consistency, whereas uncoordinated information release and unclear communication strategies increased the risk of misinformation and reputational damage.

## Discussion

This study examined operational performance during a table-top exercise of a hospital incident command structure at a single-center tertiary care university hospital, focusing on the transition from routine operations to crisis management and on identifying key operational barriers and enablers. By providing a granular operational analysis of incident command performance during activation, drawing on data from embedded participants and internal and external observers, this study offers 360° (i.e., multi-perspective insights integrating embedded participants as well as internal and external observers) triangulated insights into under-explored cognitive, communicative, and organizational processes, with the goal of informing practice and strengthening hospital crisis resilience.

The main findings, derived from nine thematic clusters shaping determinants of effective crisis response within a HICS, revealed a range of effective solutions, including (1) standardization and clear role definition, (2) structured communication with predefined pathways, (3) the use of visualization tools and checklists, (4) regular training and exercises, (5) strengthening of technical and organizational resilience, and (6) the further development of team and leadership culture. In contrast, hindering factors could be grouped into four core areas: (1) unclear structure and roles, (2) not consistently defined communication processes, (3) insufficient prioritization and information management, and (4) deficits in infrastructure and personnel.

The following section situates the findings within established resilience and emergency preparedness frameworks, juxtaposing the inductively derived framework with existing models to enable a theory-informed, deductive comparison of areas of convergence and divergence.

### Alignment of findings with existing frameworks

The identified operational clusters align closely with the nine core elements of community resilience described by Patel et al. (47) - including governance and leadership, communication, local knowledge, resources, preparedness, and social-organizational capacity - by reflecting how these domains are operationalized within a hospital incident command setting, indicating that organizational crisis management processes mirror key dimensions of resilience at the system level as in a community in general; a detailed mapping of these relationships is presented in **Table 2**.

**Table 2:**
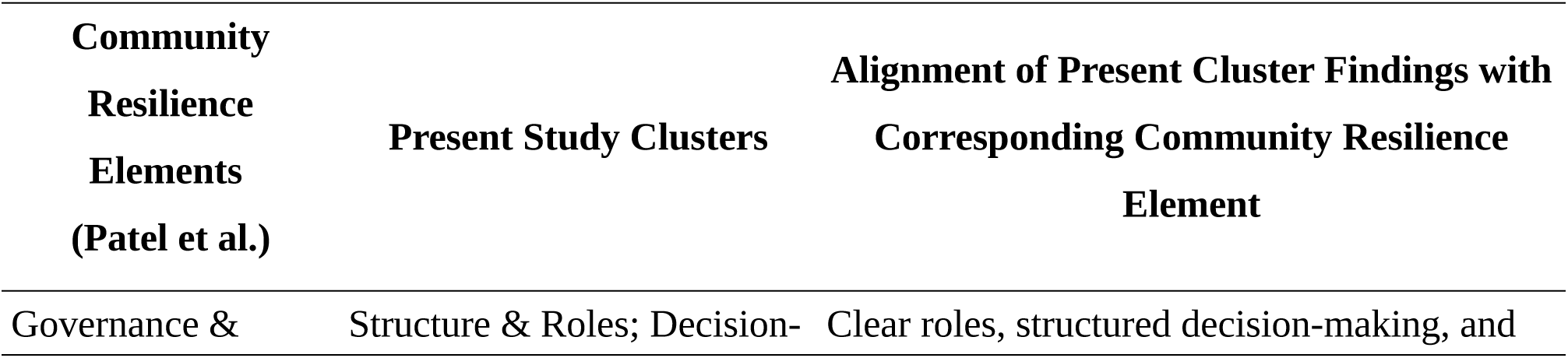
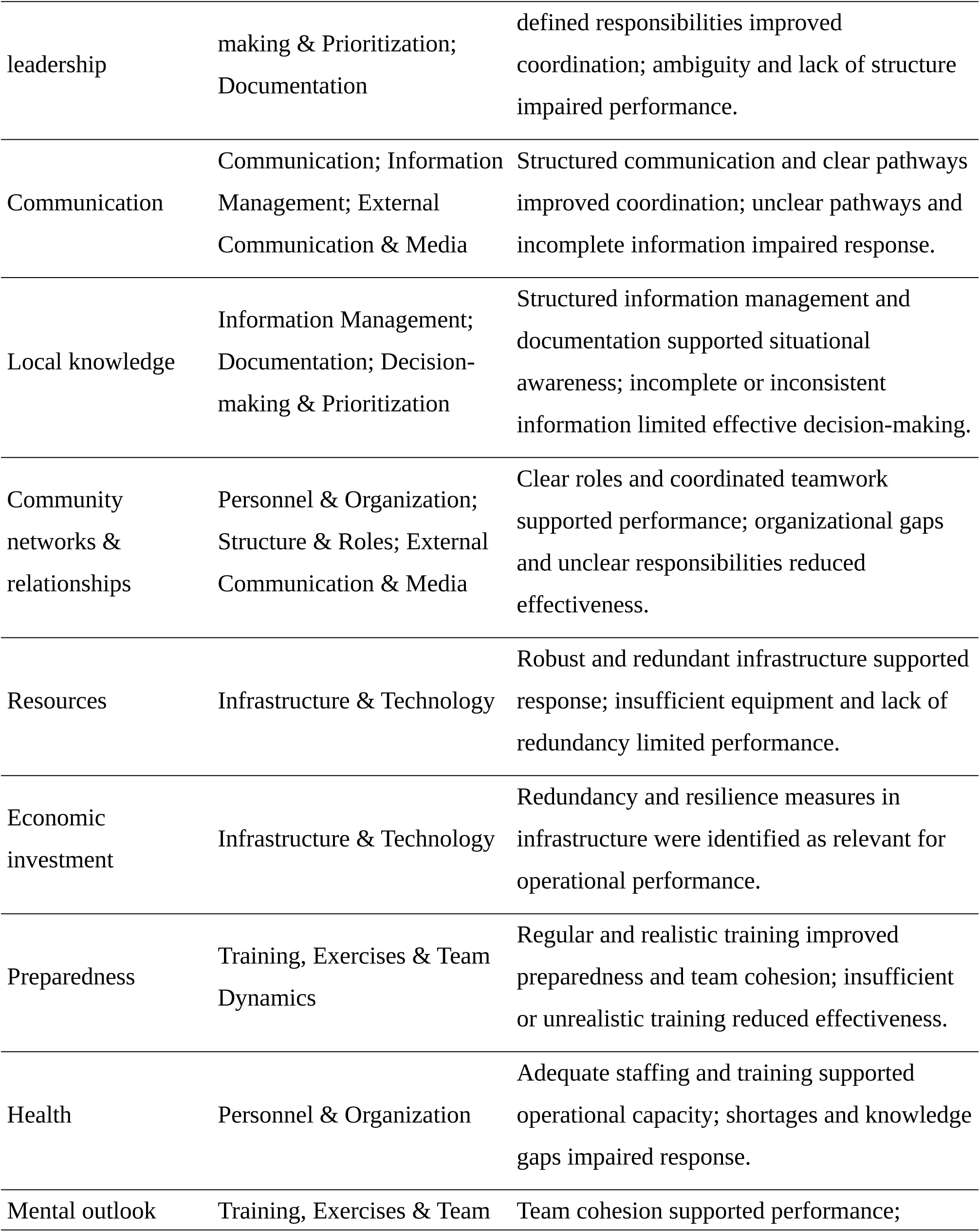
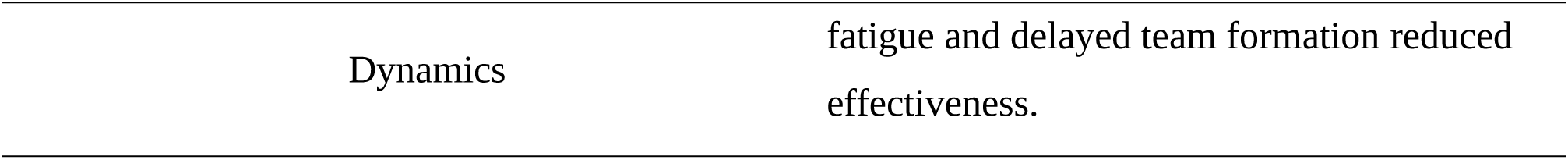
Alignment of the nine core elements of community resilience (Patel et al., 2017 (47)) with the operational clusters identified in this study that shape determinants of effective crisis response within a hospital incident command system. The table illustrates how cluster-specific findings correspond to established resilience domains at a systemic level, reflecting mechanisms also observed in community resilience contexts.

Our findings suggest that participants’ approaches to leadership and decision-making show similarities to principles described in military doctrine, including the clear articulation of leadership intent, the structured separation of information processing (progressing from facts to deductions to conclusions), and the principle of *Auftragstaktik* (often translated as mission command) (48,49) (**Table 3).** The clear intention of leadership refers to the explicit formulation of goals, priorities, and desired end states, enabling all actors to align their actions even in dynamic and uncertain environments (48). The semantic separation between fact, deduction, and conclusion reflects a disciplined cognitive process in which raw information is first collected and distinguished from interpretation, before being synthesized into decisions, thereby reducing premature action and cognitive bias (48). Finally, *Auftragstaktik* describes an intent-based leadership approach in which strategic direction is defined centrally, while execution is decentralized, allowing individuals with local expertise to adapt actions to the evolving situation (49). These three principles are deeply interrelated: Commander’s Intent provides the why, *Auftragstaktik* governs who decides and how freely, and structured intelligence processing - facts to deductions to conclusions - provides the analytical foundation on which both depend. Together they form a coherent epistemology of military leadership under uncertainty. Together, these principles appear to facilitate coordinated action under uncertainty by promoting shared situational awareness, reducing cognitive overload, and enabling flexible yet aligned decision-making. Their emergence in a civilian hospital incident command context underscores their potential relevance beyond military settings and highlights their value for strengthening crisis leadership in healthcare systems.

**Table 3.** Application of core leadership principles derived from military doctrine in hospital incident command. The table illustrates how participants applied key leadership principles commonly used in military contexts during crisis response, including clear intention of leadership, semantic separation between fact, deduction, and conclusion, and *Auftragstaktik* (mission command). Verbatim quotes from participant interviews exemplify how these principles were operationalized in practice.

| **Leadership Principle** | **Illustrative Quote(s)** | **Conceptual Integration** |
| --- | --- | --- |
| **Clear intention of leadership** | *“I have the responsibility - we will do it this way. [...] And this is what I expect” (I1).* | Defines purpose and direction |
| **Semantic separation (fact - deduction - conclusion)** | *“My conclusion is to structure the collection of information and not be tempted to jump straight into solutions and actions, but only in a second step [...] to take the time to consider what information we need and in what level of detail, then to gather that information, and only then to make decisions” (I11).* | Structures information and decision-making |
| **Auftragstaktik (mission command)** | *“[...] because you have to keep the bigger picture in mind and trust others to implement things on site, drawing on their local knowledge, in line with what has been decided at the top” (I1).*<br>*“I can’t just say I’ll work on my own tasks; if I see something where I can support, I can propose it and act on it - if it is desired”(I1).* | Defines decision authority and degree of autonomy |

Beyond community resilience and the three core military leadership principles, a range of established methodological and conceptual frameworks closely align with the nine thematic clusters identified in this study and provide complementary lenses for interpreting operational barriers and facilitators in crisis response. Besides the HICS, these include the Incident Command System (ICS), the National Incident Management System (NIMS), the Systems Engineering Initiative for Patient Safety (SEIPS) model, including SEIPS 2.0 and SEIPS 3.0, the Crisis Standards of Care framework, and the Emergency Management Program (EMP) framework. In addition, as outlined further below, integrative cross-disciplinary frameworks further enrich the analytical perspective. Specifically, the Incident Command System provides a standardized organizational structure for crisis management with five functional groups: Command (overall management), Operations (tactical execution), Planning (information collection and forecasting), Logistics (supplies, equipment, personnel support), and Finance (cost authorization and analysis) (50). This framework directly maps to several of the identified clusters including Structure and Roles, Decision-Making, and Personnel and Organization. The ICS emphasizes unified command, common terminology, and centralized coordination - principles that address the Communication and External Communication clusters (50). The National Incident Management System (NIMS) expands on ICS by providing a comprehensive framework with five core components: Preparedness, Communications and Information Management, Resource Management, Command and Management, and Ongoing Management and Maintenance (51). NIMS creates a common operating picture across federal, state, and local levels, ensuring consistency in incident information and decision-making. (51). This framework encompasses the present Information Management, Infrastructure and Technology, and Training clusters. The HICS adapts ICS specifically for healthcare facilities and operates through five major sections that parallel the thematic clusters identified in this study (51). HICS’ effectiveness relies on staff familiarity through training, routine exercises, and activation for incidents requiring rapid multifaceted response - directly addressing Training, Exercises, and Team Dynamics cluster (51). The phenomena observed in the Training, Exercises, and Team Dynamics cluster can be interpreted within the well-described “panic–then-forget cycle” and the transient “Walker dip,” suggesting that initial performance deficits during incident command activation are not signs of failure but expected transitional phenomena, underscoring the need for maintenance of skills through regular training and institutional memory to prevent recurrent loss of preparedness between crises (52,53). The SEIPS model (Systems Engineering Initiative for Patient Safety) offers a sociotechnical systems approach examining how people, tools, tasks, and organizational environments interact to influence processes and outcomes (51,54,55). SEIPS 2.0 and 3.0 incorporate concepts of configuration, engagement, and adaptation that explain how dynamic systems evolve during crises (55). This framework has been successfully applied to identify both barriers and facilitators in healthcare settings, making it particularly relevant for analyzing the present operational factors (56,57). Additional relevant frameworks include the Crisis Standards of Care model, which distinguishes conventional, contingency, and crisis surge responses based on resource availability and decision-making shifts from patient-centered to population-centered care (58). The Emergency Management Program (EMP) framework structures crisis management into four phases-preparedness, mitigation, response, and recovery-and encompasses key components such as communication, resource management, safety, and staff roles (51,59,60). Integrated approaches that combine and align multiple frameworks are gaining prominence. Iserson proposes combining sensemaking, recognition-primed decision-making, the Cynefin framework, complexity theory, edge-of-chaos theory, and ICS to enhance decision-making and operational effectiveness during disasters (61). Likewise, a scoping review by Sasie et al. identified four core domains for evaluating public health emergency management: overall implementation level, individual factors (capacity building, resources, engagement), organizational factors (information sharing, planning, coordination, infrastructure), and overarching factors (workforce, funding, governance) (62).

In general, our nine thematic clusters align conceptually with established frameworks, particularly HICS and SEIPS, which provide structured approaches to identifying and addressing operational barriers and facilitators. The SEIPS model is particularly useful for analyzing interactions between clusters, while HICS offers a structured framework for operational implementation. However, although these models define relevant domains and structures, they offer limited insight into how processes unfold in real time. This study addresses this gap by examining what actually happens within HICS during activation. Rather than focusing solely on formal structures or outcome metrics, it opens the “black box” of incident command by exploring the internal dynamics that shape real-time crisis response. Methodologically, the present report introduces a narrative-phenomenological, triangulated design to a field that has widely relied on instrumental and quantitative approaches, thereby enabling a richer understanding of lived experiences and operational processes. Furthermore, the study repositions the table-top exercise as an analytical environment, allowing for the isolation of cognitive processes, the observation of communication patterns, and the examination of decision-making mechanisms under controlled yet realistic conditions. In doing so, it generates micro-level, process-based insights into coordination, communication, and decision-making. Finally, it provides empirical grounding for how incident command structures function in practice, where they may break down, and where adaptation is required, thereby offering a more nuanced understanding of their real-world performance in complex and dynamic crisis settings.

### Limitations, bias mitigation strategies, generalizability, and directions for future research

Several important limitations including potential sources ob bias should be considered when interpreting the findings of this study. First, a single-site design at a large university hospital may limit transferability to settings with different structures, resources, or levels of preparedness. Second, the table-top exercise, while enabling focused analysis of cognitive, communicative, and decision-making processes, cannot fully replicate the complexity and time pressure of real-world emergencies. Third, the qualitative design and reliance on self-reported data may introduce subjective bias, including recall and social desirability effects. Fourth, the sample was restricted to participants within a specific training context, potentially limiting the diversity of perspectives. Sixth, voluntary participation may result in selection bias.

These limitations were mitigated through methodological triangulation, integrating semi-structured interviews, participant observation, and document analysis to enhance robustness. Data collection and analysis followed an iterative, narrative-phenomenological approach (Creswell), including significant statements, horizontalization, and clustering of meaning, supported by transparent reporting, reflexivity, and adherence to established qualitative standards. In addition, the study combined perspectives from embedded researchers - providing in-depth contextual insight into processes during the exercise - with non-embedded observers, both from the local institution and from external institutions, contributing independent viewpoints and reducing the risk of single-institution or role-specific bias. This combination of insider and outsider perspectives strengthened the credibility and interpretive robustness of the findings.

The identified barriers and enablers relate to core processes-communication, coordination, and decision-making-central to HICS in general and are embedded in widely used frameworks, supporting transferability to comparable settings. This report focuses on operational enablers and barriers, while a separate report (manuscript in preparation), based on a Grounded Theory approach, develops a theoretical understanding of leadership orchestration processes using the same table-top exercise.

Within these constraints, the findings may be transferable to comparable hospital settings, particularly those operating within similar structured command systems.

Future research should extend this work through multi-center studies across diverse healthcare systems, incorporating full-scale simulations and real-world incident data. Mixed-methods designs linking qualitative insights to quantitative performance metrics, as well as studies on leadership dynamics and targeted interventions, may further advance understanding and practical implementation.

## Conclusion

Preparedness frameworks are necessary but insufficient as stand-alone approaches, as their operational execution ultimately determines real-world performance. This is reflected in recurring deficits such as unclear roles and structures, inconsistent communication, weak prioritization and information management, and gaps in infrastructure and personnel. At the same time, the findings indicate that a small number of standardized cognitive and organizational practices can substantially enhance crisis performance under uncertainty, including clear role separation and predefined responsibilities, explicitly communicated leadership intent to align actions and priorities, structured closed-loop communication, explicit decision cycles from information gathering to structuring to decision-making, the use of checklists, visualization, and central information management, as well as the acceptance of rapid “80% decisions.” Embedded within this, the principle of *Auftragstaktik* (mission command) - combining clear strategic intent with decentralized execution - further enables adaptive and coordinated action in dynamic environments. Finally, as hospital incident command represents a microcosm of system resilience, strengthening these structures is not merely an internal optimization, but a key lever for achieving whole-of-system resilience in crises.

## Data Availability

All data produced in the present study are available upon reasonable request to the authors

## Acknowledgments

The authors thank all participants for their time, commitment, and valuable contributions to this study. We also acknowledge the University Hospital Heidelberg for its strong commitment to strengthening resilience and crisis preparedness, which made this project possible.

## Data Availability Statement

The datasets generated and analyzed during the current study are not publicly available due to the risk of indirect participant identification in a small, single-center qualitative dataset containing role-specific and context-rich information. In addition, the data includes sensitive organizational insights that are subject to institutional confidentiality. Data sharing is therefore restricted by ethical approval and applicable data protection regulations (GDPR). De-identified excerpts supporting the findings may be made available from the corresponding author on reasonable request, subject to appropriate safeguards.

## Author Contributions (CrediT)

Conceptualization: EP, JK, MF, MR

Methodology: JK, MF, MR

Data collection: HS, JK, KB, MF, MR

Analysis: FW, HS, JK, KB, MF, MR

Writing original draft: MR

Review & editing: EP, FW, HS, JK, KB, MF, MR, KF, PG

## References

1. Watts N, Amann M, Ayeb-Karlsson S, Belesova K, Bouley T, Boykoff M, et al. The Lancet Countdown on health and climate change: from 25 years of inaction to a global transformation for public health. Lancet. 2018 Feb 10;391(10120):581–630. doi:10.1016/S0140-6736(17)32464-9 PubMed PMID: 29096948.

2. Traore T, Shanks S, Haider N, Ahmed K, Jain V, Rüegg SR, et al. How prepared is the world? Identifying weaknesses in existing assessment frameworks for global health security through a One Health approach. Lancet. 2023 Feb 25;401(10377):673–87. doi:10.1016/S0140-6736(22)01589-6 PubMed PMID: 36682374.

3. von der Forst M, Dietrich M, Schmitt FCF, Popp E, Ries M. Perennial disaster patterns in Central Europe since 2000 and implications for hospital preparedness planning - a cross-sectional analysis. Sci Rep. 2025 Jan 3;15(1):620. doi:10.1038/s41598-024-84223-4 PubMed PMID: 39753701; PubMed Central PMCID: PMC11698994.

4. Klokman VW, Barten DG, Peters NALR, Versteegen MGJ, Wijnands JJJ, van Osch FHM, et al. A scoping review of internal hospital crises and disasters in the Netherlands, 2000-2020. PLoS One. 2021;16(4):e0250551. doi:10.1371/journal.pone.0250551 PubMed PMID: 33901248; PubMed Central PMCID: PMC8075216.

5. Hassan EM, Mahmoud H. Healthcare and education networks interaction as an indicator of social services stability following natural disasters. Sci Rep. 2021 Jan 18;11(1):1664. doi:10.1038/s41598-021-81130-w PubMed PMID: 33462303; PubMed Central PMCID: PMC7814048.

6. Gogalniceanu P. Hybrid threats require hybrid solutions: A roadmap for healthcare security. Health Policy. 2026 Apr;166:105565. doi:10.1016/j.healthpol.2026.105565 PubMed PMID: 41570631.

7. Hugelius K, Becker J, Adolfsson A. Five Challenges When Managing Mass Casualty or Disaster Situations: A Review Study. Int J Environ Res Public Health. 2020 Apr 28;17(9):3068. doi:10.3390/ijerph17093068 PubMed PMID: 32354076; PubMed Central PMCID: PMC7246560.

8. Schulze C, Welker A, Kühn A, Schwertz R, Otto B, Moraldo L, et al. Public Health Leadership in a VUCA World Environment: Lessons Learned during the Regional Emergency Rollout of SARS-CoV-2 Vaccinations in Heidelberg, Germany, during the COVID-19 Pandemic. Vaccines (Basel). 2021 Aug 11;9(8):887. doi:10.3390/vaccines9080887 PubMed PMID: 34452012; PubMed Central PMCID: PMC8402600.

9. Kaye AD, Cornett EM, Kallurkar A, Colontonio MM, Chandler D, Mosieri C, et al. Framework for creating an incident command center during crises. Best Pract Res Clin Anaesthesiol. 2021 Oct;35(3):377–88. doi:10.1016/j.bpa.2020.11.008 PubMed PMID: 34511226; PubMed Central PMCID: PMC8428470.

10. Joint Commission. National Performance Goals^TM^ Effective January 2026 for the Hospital Program [Internet]. 2025 [cited 2026 Mar 30]. Available from: https://digitalassets.jointcommission.org/api/public/content/9ca80055182b4274842a5780a94f2c82

11. Klinger I, Heckel M, Shahda S, Krisen U, Stellmacher S, Kurkowski S, et al. [COVID-19 pandemic response teams: organization, competencies, and challenges-understanding and using structural realities]. Bundesgesundheitsblatt Gesundheitsforschung Gesundheitsschutz. 2022 Jun;65(6):650–7. doi:10.1007/s00103-022-03542-x PubMed PMID: 35503572; PubMed Central PMCID: PMC9063247.

12. AFKzV. Feuerwehr-Dienstvorschrift 100, Ausgabe: März 1999, Führung und Leitung im Einsatz [Internet]. 1999. Available from: https://www.lfs-bw.de/fileadmin/LFS-BW/themen/gesetze_vorschriften/fwdv/dokumente/FwDV_100.pdf

13. von der Forst M, Popp E, Schaefer H, Bikowski K, Ries M, Kuellenberg J. Exploring the Continental Staff System as a Framework for the Hospital Incident Command – Protocol for a Qualitative Grounded Theory Study [Internet]. medRxiv; 2025 [cited 2026 Mar 30]. p. 2025.05.01.25326786. Available from: https://www.medrxiv.org/content/10.1101/2025.05.01.25326786v1 doi:10.1101/2025.05.01.25326786

14. Geiger M, Neuner S, Bodur ME, Fekete A. [Failure of critical infrastructures in German hospitals : Online survey on crisis management structures and exercises in hospitals]. Anaesthesiologie. 2026 Mar 27. doi:10.1007/s00101-026-01664-4 PubMed PMID: 41894032.

15. WHO. International Health Regulations (2005) – Third edition [Internet]. 2005 [cited 2026 Mar 30]. Available from: https://www.who.int/publications/i/item/9789241580496

16. WHO. Essential public health functions, health systems and health security [Internet]. 2018 [cited 2026 Mar 30]. Available from: https://www.who.int/publications/i/item/9789241514088

17. WHO. Rapid hospital readiness checklist: Interim Guidance [Internet]. 2020 [cited 2026 Mar 30]. Available from: https://www.who.int/publications/i/item/WHO-2019-nCoV-hospital-readiness-checklist-2020.1

18. WHO. Hospital emergency response checklist [Internet]. 2011 [cited 2026 Mar 30]. Available from: https://www.who.int/publications/i/item/hospital-emergency-response-checklist

19. BBK. Handbuch Krankenhausalarm- und -einsatzplanung (KAEP) [Internet]. 2020 [cited 2026 Mar 30]. Available from: https://www.bbk.bund.de/SharedDocs/Downloads/DE/Mediathek/Publikationen/Gesundheit/KAEP/handbuch-kaep.pdf?blob=publicationFile&v=20

20. BBK. Handbuch KAEP Einleger Grafik [Internet]. 2020 [cited 2026 Mar 30]. Available from: https://www.bbk.bund.de/SharedDocs/Downloads/DE/Mediathek/Publikationen/Gesundheit/handbuch-kaep-einleger-grafik.pdf?blob=publicationFile&v=9

21. DAKEP. Anforderungen an die DAKEP-Zertifizierung der Krankenhausalarm- und Einsatzplanung (KAEP) [Internet]. 2020. Available from: https://www.dakep-active.de/app/download/14554554932/AnfordergKatalog_Zert.pdf

22. Sozialministerium Baden-Württemberg. Rahmenplan zur Krankenhausalarm- und Einsatzplanung Baden-Württemberg [Internet]. 2025 [cited 2026 Mar 30]. Available from: https://sozialministerium.baden-wuerttemberg.de/fileadmin/redaktion/m-sm/intern/downloads/Publikationen/Rahmen-KAEP-BW_2025.pdf

23. Ries M. Towards Better Preparedness for Future Catastrophes: Lessons Learned from Civil-Military Pandemic Response [Internet]. Heidelberg: heiBOOKS; 2024. Available from: 10.11588/heibooks.1331 https://doi.org/10.11588/heibooks.1331

24. Hayirli TC, Kuznetsova M, Biddinger PD, Bambury EA, Atkinson MK. Formal and informal hospital emergency management practices: managing for safety and performance amid crisis. Int J Qual Health Care. 2024 Jul 31;36(3):mzae069. doi:10.1093/intqhc/mzae069 PubMed PMID: 38988191.

25. Verheul ML, Dückers ML. Defining and Operationalizing Disaster Preparedness in Hospitals: A Systematic Literature Review. Prehosp Disaster Med. 2020 Feb;35(1):61–8. doi:10.1017/S1049023X19005181 PubMed PMID: 31826788.

26. Williams J, Nocera M, Casteel C. The effectiveness of disaster training for health care workers: a systematic review. Ann Emerg Med. 2008 Sep;52(3):211–22, 222.e1-2. doi:10.1016/j.annemergmed.2007. 09.030 PubMed PMID: 18069087.

27. Hsu EB, Jenckes MW, Catlett CL, Robinson KA, Feuerstein C, Cosgrove SE, et al. Effectiveness of hospital staff mass-casualty incident training methods: a systematic literature review. Prehosp Disaster Med. 2004;19(3):191–9. doi:10.1017/s1049023x00001771 PubMed PMID: 15571194.

28. Merriel A, Ficquet J, Barnard K, Kunutsor SK, Soar J, Lenguerrand E, et al. The effects of interactive training of healthcare providers on the management of life-threatening emergencies in hospital. Cochrane Database Syst Rev. 2019 Sep 24;9(9):CD012177. doi:10.1002/14651858.CD012177.pub2 PubMed PMID: 31549741; PubMed Central PMCID: PMC6757513.

29. von der Forst M, Germann BJ, Schaefer H, Salg GA, Weigand MA, Schmitt FCF, et al. Impact of a full-scale mass casualty exercise on hospital staff and implications for future preparedness – A pre-post study. Progress in Disaster Science. 2025 Dec 1;28:100478. doi:10.1016/j.pdisas.2025.100478

30. von der Forst M, Schaefer H, Mohr S, Kenngott HG, Farjallah E, Huck M, et al. Improving preparedness for mass casualty incidents in hospitals: insights from a large-scale simulation exercise with geotracking and validated questionnaires. BMC Emerg Med. 2026 Mar 9. doi:10.1186/s12873-026-01527-6 PubMed PMID: 41803710.

31. von der Forst M. The Service Regulation DV 100 as an Organizational Structure for Establishing the Hospital Incident Command System [Clinical trial registration] [Internet]. clinicaltrials.gov; 2025 [cited 2026 Mar 30]. Available from: https://clinicaltrials.gov/study/NCT06913010

32. Creswell JW. Narrative Research. In: Qualitative Inquiry & Research Design: Choosing Among Five Approaches. Washington, DC, USA: Sage; 2012. p. 70–6.

33. Creswell JW. Phenomenological Research. In: Qualitative Inquiry & Research Design: Choosing Among Five Approaches. Washington, DC, USA: Sage; 2012. p. 76–83.

34. Charmaz K. Constructivist grounded theory. The Journal of Positive Psychology. 2017 May 4;12(3):299–300. doi:10.1080/17439760.2016.1262612

35. Mey G, Mruck K. Grounded-Theory-Methodologie. In: Handbuch Qualitative Forschung in der Psychologie. 2nd ed. 2020. p. 513–35.

36. O’Brien BC, Harris IB, Beckman TJ, Reed DA, Cook DA. Standards for Reporting Qualitative Research: A Synthesis of Recommendations. Academic Medicine. 2014 Sep;89(9):1245–51. doi:10.1097/ACM.0000000000000388

37. Elm E von, Altman DG, Egger M, Pocock SJ, Gøtzsche PC, Vandenbroucke JP, et al. The Strengthening the Reporting of Observational Studies in Epidemiology (STROBE) Statement: Guidelines for Reporting Observational Studies. PLOS Medicine. 2007 Oct 16;4(10):e296. doi:10.1371/journal.pmed.0040296

38. Kruse J. Qualitative Interviewforschung: ein integrativer Ansatz. 2., überarbeitete und ergänzte Auflage. Weinheim ; Basel: Beltz Juventa; 2015. 707 Seiten. (Grundlagentexte Methoden).

39. Ries M. The COVID-19 Infodemic: Mechanism, Impact, and Counter-Measures—A Review of Reviews. Sustainability. 2022 Jan;14(5):2605. doi:10.3390/su14052605

40. Ries M. Global key concepts of civil-military cooperation for disaster management in the COVID-19 pandemic—A qualitative phenomenological scoping review. Front Public Health. 2022 Sep 15;10. doi:10.3389/fpubh.2022.975667

41. Ries M. Resilienz von Kindern und Jugendlichen bei Katastrophen stärken. Forum Marsilius-Kolleg. 2025 Mar 4;25. doi:10.11588/fmk.2025.25.109299

42. Curtain C. QualCoder 3.3 [Computer software] [Internet]. 2023 [cited 2026 Mar 31]. Available from: https://github.com/ccbogel/QualCoder/releases/tag/3.3

43. Linux Mint. Linux Mint 21.3 “Virginia” - Linux Mint [Internet]. [cited 2026 Mar 31]. Available from: https://linuxmint.com/edition.php?id=311

44. Williams T. Minder 2.0.5 [Vala] [Internet]. 2026 [cited 2026 Apr 6]. Available from: https://github.com/phase1geo/Minder

45. R Core Team. R: A language and environment for statistical computing. R Foundation for Statistical Computing, Vienna, Austria. [Internet]. 2026 [cited 2026 Apr 6]. Available from: https://www.r-project.org/

46. Posit team. Posit RStudio: Integrated Development for R. Posit Software, PBC, Boston, MA. [Internet]. 2026 [cited 2026 Apr 6]. Available from: https://www.posit.co/

47. Patel SS, Rogers MB, Amlôt R, Rubin GJ. What Do We Mean by “Community Resilience”? A Systematic Literature Review of How It Is Defined in the Literature. PLoS Curr. 2017 Feb 1;9:ecurrents.dis.db775aff25efc5ac4f0660ad9c9f7db2. doi:10.1371/currents.dis.db775aff25efc5ac4f0660ad9c9f7db2 PubMed PMID: 29188132; PubMed Central PMCID: PMC5693357.

48. CCOE. CIMIC COE [Internet]. 2026 [cited 2026 Mar 31]. 5.2. Tactical Planning Process. Available from: https://www.cimic-coe.org/handbook-entries/welcome-to-the-cimic-handbook/v-cimic-contribution-to-the-planning-processes/5-2-tactical-planning-process/

49. Sjøgren S, Nilsson N. Multinational Mission Command: From Paper to Practice in NATO. Scandinavian Journal of Military Studies. 2025 Apr 16;8(1). doi:10.31374/sjms.329

50. Waeckerle JF. Disaster planning and response. N Engl J Med. 1991 Mar 21;324(12):815–21. doi:10.1056/NEJM199103213241206 PubMed PMID: 1997854.

51. Banach DB, Johnston BL, Al-Zubeidi D, Bartlett AH, Bleasdale SC, Deloney VM, et al. Outbreak Response and Incident Management: SHEA Guidance and Resources for Healthcare Epidemiologists in United States Acute-Care Hospitals. Infect Control Hosp Epidemiol. 2017 Dec;38(12):1393–419. doi:10.1017/ice.2017.212 PubMed PMID: 29187263; PubMed Central PMCID: PMC7113030.

52. WHO. Sustainable preparedness for health security and resilience: Adopting a whole-of-society approach and breaking the “panic-then-forget” cycle [Internet]. World Health Organization; 2020 [cited 2026 Mar 31]. Available from: https://www.jstor.org/stable/resrep55479

53. Walker AJ. The ‘Walker dip.’ Journal of The Royal Naval Medical Service. 2018 Dec 21;104(3). doi:10.1136/jrnms-104-173

54. Carayon P, Wetterneck TB, Rivera-Rodriguez AJ, Hundt AS, Hoonakker P, Holden R, et al. Human factors systems approach to healthcare quality and patient safety. Appl Ergon. 2014 Jan;45(1):14–25. doi:10.1016/j.apergo.2013.04.023 PubMed PMID: 23845724; PubMed Central PMCID: PMC3795965.

55. Holden RJ, Carayon P, Gurses AP, Hoonakker P, Hundt AS, Ozok AA, et al. SEIPS 2.0: a human factors framework for studying and improving the work of healthcare professionals and patients. Ergonomics. 2013;56(11):1669–86. doi:10.1080/00140139.2013.838643 PubMed PMID: 24088063; PubMed Central PMCID: PMC3835697.

56. Leon C, Hogan H, Jani YH. Seeking systems-based facilitators of safety and healthcare resilience: a thematic review of incident reports. Int J Qual Health Care. 2024 Jul 9;36(3):mzae057. doi:10.1093/intqhc/mzae057 PubMed PMID: 38915190; PubMed Central PMCID: PMC11233260.

57. Berman L, Kavalier M, Gelana B, Tesfaw G, Siraj D, Shirley D, et al. Utilizing the SEIPS model to guide hand hygiene interventions at a tertiary hospital in Ethiopia. PLOS ONE. 2021 Oct 28;16(10):e0258662. doi:10.1371/journal.pone.0258662

58. Hick JL, Einav S, Hanfling D, Kissoon N, Dichter JR, Devereaux AV, et al. Surge capacity principles: care of the critically ill and injured during pandemics and disasters: CHEST consensus statement. Chest. 2014 Oct;146(4 Suppl):e1S–e16S. doi:10.1378/chest.14-0733 PubMed PMID: 25144334.

59. Joint Commission. R3 Report Issue 39: New and Revised Emergency Management Standards for Ambulatory Care Programs. [Internet]. 2023 [cited 2026 Mar 31]. Available from: https://digitalassets.jointcommission.org/api/public/content/4406f791a9464da29ac9bc6d6ec69953

60. Joint Commission. R3 Report Issue 46: New and Revised Emergency Management Standards for Nursing Care Centers. [Internet]. 2023 [cited 2026 Mar 31]. Available from: https://digitalassets.jointcommission.org/api/public/content/4406f791a9464da29ac9bc6d6ec69953

61. Iserson KV. Integrating Disaster Response Tools for Clinical Leadership. West J Emerg Med. 2025 Jan;26(1):30–9. doi:10.5811/westjem.35390 PubMed PMID: 39918139; PubMed Central PMCID: PMC11908526.

62. Sasie SD, Ayano G, Van Zuylen P, Aragaw FM, Darebo TD, Guerrero-Torres L, et al. Developing a comprehensive framework for evaluating public health emergency management program implementation: A scoping review. Public Health. 2025 Feb;239:22–31. doi:10.1016/j.puhe.2024.12.012 PubMed PMID: 39721141.

